# Rapid prototyping and clinical testing of a reusable face shield for health care workers responding to the COVID-19 pandemic

**DOI:** 10.1101/2020.04.11.20061960

**Authors:** Arash Mostaghimi, M.-J. Antonini, Deborah Plana, Philip D. Anderson, Brandon Beller, Edward W. Boyer, Amber Fannin, Jacob Freake, Richard Oakley, Michael S. Sinha, Leanne Smith, Christopher Van, Helen Yang, Peter K. Sorger, Nicole R. LeBoeuf, Sherry H. Yu

## Abstract

Due to supply chain disruption, the COVID-19 pandemic has caused severe shortages in personal protective equipment (PPE) for health care professionals. Local fabrication based on 3D printing is one way to address this challenge, particularly in the case of simple products such as protective face shields. As a consequence, many public domain designs for face shields have become available. No clear path exists, however, for introducing a locally fabricated and unapproved product into a clinical setting. In a US health care setting, face shields are regulated by the Food and Drug Administration (FDA); similar policies exist in other countries. We describe a research protocol under which rapid iteration on an existing design, coupled with clinical feedback and real-world testing in an emergency department, allowed a face shield to be implemented by the members of the incident command team at a major academic medical center. We describe our design and testing process and provide an overview of regulatory considerations associated with fabrication and testing of face shields and related products. All designs, materials used, testing protocols, and survey results are reported in full to facilitate the execution of similar face shield efforts in other clinical settings. Our work serves as a case study for development of a robust local response to pandemics and other health care emergencies, with implications for healthcare professionals, hospital administrators, regulatory agencies and concerned citizens.

## Introduction

In the face of a rapidly expanding COVID-19 pandemic in the winter of 2019 and spring of 2020, severe shortages have emerged in personal protective equipment (PPE), putting both health care professionals and patients at increased risk of infection. The origins of these shortages are varied but reflect the fragility of medical supply chains in which relatively few international vendors dominate many critical medical product areas. Because many hospitals use just-in-time inventory management, supply chain problems rapidly deplete hospital supplies and prevent restocking from traditional vendors. Faced with this crisis, many caregivers and medical centers have turned to local fabricators to see if they can provide replacements for products such as face shields, filtering respirators, and even ventilators. The substitution of conventionally sourced products with non-traditional local products is made feasible by rapid expansion in inexpensive additive manufacturing capabilities (“3D printing”) by small business and hobbyist (“maker”) communities. Computer-aided design (CAD) software has also become widely available, making it possible to share designs in public forums, including the NIH 3D Print Exchange^1^. This has resulted in dozens of open-sourced designs, online videos, and blogs dedicated to fabricating different types of PPE.

This article describes the local fabrication and testing of a face shield, one of the simpler types of PPE in terms of design and regulation, from prototyping through clinical testing and adoption by the members of incident command^2^ of a major hospital system. We discuss how hospital systems can most effectively test and make use of alternative or innovative products in the face of life-threatening disease while ensuring staff and patient safety. This is a critical issue. Multiple projects led by small companies and citizens are asking how the PPE they have fabricated can be provided to hospitals in need - the ‘last mile’ of the supply chain. We review the regulatory guidance on this issue and explore the life cycle of a product implemented in a crisis situation, including whether a product adopted in crisis should remain in inventory after the crises has passed.

Face shields are used in hospitals for infection control,^3^ and are also required PPE in many research and industrial settings. Although they are simple appearing devices they are subject to regulation. In the US, the ANSI/ISEA Z.87.1-2015 standard specifies nearly twenty required physical features of a face shield and testing requirements for visual resolving power, resistance to high-velocity impacts, and protection from droplets and splashes. Similar standards exist in Europe and other countries. The need for such standards is obvious given that defective face shields can expose users, particularly those in industry, to serious and life-threatening injuries (e.g., in welding). In a health care setting, face shields are categorized as Class I medical devices, the least-regulated FDA category.

Typically, a manufacturer passes an ANSI/ISEA Z.87.1-2015 certification and then notifies the FDA of compliance. The FDA (and in some cases the CDC) maintain a list of approved products^4^. Unlike more complex medical products, a 510(k) filing is not required; 510(k) filings require a demonstration that a device is substantially equivalent to an existing, legally marketed device. Except in rare circumstances, local manufacturers, maker communities and hospitals are unlikely to have the necessary expertise to test face shields to ANSI/ISEA Z.87.1-2015. However, as of April 2020, the FDA has provided guidance that it “does not intend to object to individuals’ distribution and use of improvised PPE when no alternatives, such as FDA-cleared masks or respirators, are available.” ^5^ This provides a regulatory framework in which to use locally fabricated (“improvised”) PPE, but it does not address a critical question: how can these devices be introduced into the hospital supply chain in a rational, safe and controlled manner?

Options for face shields being pursued by individual citizens, nonprofit institutions, academic medical centers, and small and large-scale manufacturers include flat plastic shields that can be rapidly assembled by users, three-part designs consisting of a shield, elastic headband, and brow foam, which are being manually assembled by volunteers across the country, and 3D-printed shields including the Prusa design and its derivatives (**Table 1**). These designs have been introduced with different use cases in mind. Unlike industrial face shields, which can be expensive and are often used for extended periods of time, the vast majority of medical face shields are low-cost and intended to be discarded after a single use. Some non-regulated designs (e.g. from Prusa^6^) are intended for multiple uses and are potentially superior in fit and function to regulated disposable face shields. As a practical matter, at a time when PPE is in extremely short supply, even lower-quality face shields are unlikely to be discarded after a single use. This raises questions about procedures for face shield sterilization, which also requires testing and evaluation.

**Table 1:** Examples of ongoing, non-traditional face-shield fabrication designs and specific efforts.

| Face shield design description | Links to specific design efforts |
| --- | --- |
| Flat plastic face shields which can be rapidly assembled by users | https://project-manus.mit.edu/fs<br>https://open-face-website.now.sh/ |
| 3-part machine-less face-shield requiring volunteer assembly | https://making.engr.wisc.edu/shield/ |
| 3D-printed face shields requiring manufacturer assembly | https://www.prusaprinters.org/prints/25857<br>https://www.protohaven.org/proto-shield/<br>https://3dprint.nih.gov/discover/3dpx-013359 |

In this paper we describe the production and implementation of a 3D printed face shield (modified from the Prusa design^6^ developed in the Czech Republic) and its introduction into the Brigham and Women’s Hospital (BWH), a major US academic medical center. In conjunction with the members of BWH Incident Command, we obtained user feedback from surveyed emergency department staff under an Institutional Review Board (IRB)^7^-approved protocol. We also describe modifications to the design to prepare it for large-scale manufacturing through industry partnerships. The use of a research protocol made it possible to introduce an untested device, and ready it for deployment, in advance of FDA guidance and in a manner that greatly increased the confidence of hospital leadership. All of the designs and protocols generated through this effort are being freely shared for reuse and improvement, and the results for our testing at the BWH emergency department are reported in full to facilitate the execution of similar face shield efforts in other clinical settings. We anticipate that this work will provide a framework for the design and implementation of similar approaches to PPE manufacturing for current and future shortages.

## Methods

### Initial Design and Serial Prototyping

We recruited a team of five clinicians, including physicians specializing in internal medicine, infectious disease, emergency medicine and dermatology who worked in tandem with a safety officer to solicit feedback and serially prototype potential face shield designs. Starting with the open source Prusa-design, we iteratively modified, 3D-printed, and obtained clinician feedback on specific features (described in **Table 2**). Four design iterations led to consensus on a design with acceptable fit, comfort, degree of protection and use of readily-available materials. This model was officially evaluated by infection control and safety officers and approved for clinical testing. The final model, the BWH/PanFab Mk1.0 face shield (henceforth the PanFab face shield), is composed of five components: (i) a transparent visor made of biaxially-oriented polyethylene terephthalate (BoPET, also known as Mylar; LEVOSHUA brand from Amazon.com), (ii-iii) a 3D printed headband and bottom reinforcement bracket made of polylactic acid (PLA, 1.75mm diameter, Hatchbox), (iv a hook and loop strap (VELCRO® Brand ONE-WRAP; Manchester NH) and (v) a foam pad made of ethylene-vinyl acetate (EVA 6mm - unknown manufacturer, donated) for added comfort.

**Table 2:** Examples of original design features, clinical feedback for improvement, and final product.

| Original Prusa Design | Clinical Feedback for Design Improvement | Final Design |
| --- | --- | --- |
| Open gap between outer face shield envelope and user | Limited fluid protection on top of visor when performing procedures (e.g., intubation) | Added fin on top of the prototype headband and additional plastic lip to retain fluid and prevent it from obstructing face shield view |
| Single attachment point for face shield strap | Difficulty attaching strap and suboptimal fit for different face types | Used hook and loop Velcro™ to adapt each visor to individual users. |
| 240 mm width and 240 mm length for face shield outer envelope dimensions | Original length not sufficiently protective for all user facial lengths and height | Outer envelope length modified to be 240 mm wide and 305 mm long without obstructing hearing or access to ears for stethoscope |
| Anchor point for straps placed lateral to the headband | Shield uncomfortable to wear for an extended time | Anchor points for hook and loop strap placed in-line with the headbands, reducing tightness |

### Design and printing of the headband and bottom reinforcement bracket

Based on the Prusa RC2 open-source model, a 3D mesh model was imported into Fusion 360 (Autodesk, V2.0.7830) software and converted into a solid body for editing using the boundary representation (BRep). The design was then modified iteratively based on clinician feedback (summarized in **Table 2**). For each prototype, the model was exported into .STL format before being imported into the open-source 3D-printing software CURA (Ultimaker), where it was sliced using the following parameters: 0.2mm layer height, 15% gyroid infill. Following slicing, the printer-specific g-code was sent to fused filament fabrication (FFF) 3d-printers (Ender 3 Pro, Creality). PLA was 3D printed at 90mm/s, the overall material volume used, and part envelope were 53.2 × 103 mm3 and 215 × 152 × 50 mm for the headband, and 4260 mm3 and 120 × 30 × 13mm for the reinforcement bracket. These parameters were optimized to decrease print time and material, while retaining functionality. **Supplementary Material 2** includes design .STL files and **Supplementary Material 3** describes associated material considerations.

### Design and cutting of the transparent visor and the foam pad

The transparent visor was designed using InkScape software to match the pegs of the 3D-printed headband. The visor was 240 mm long and 305 mm wide, ensuring that the user’s face would be fully covered without obstructing hearing. The model was outputed into .DXF format and then laser cut from 0.007” BoPET using a GlowForge or GlowForge plus laser cutter (1 pass, speed setting: 500 mm/sec, 40% power, focus height: 0.178 mm). GlowForge and GlowForge plus maximum laser powers were 40 W and 45 W respectively. The foam pad was designed in InkScape software before being outputted into .DXF format and laser cut from 6mm EVA foam (1 pass, speed setting: 155 mm/sec, 30% power, focus height: 6.13 mm). The overall dimensions of the foam were 6 mm in thickness, 20 mm in width, and 190 mm in length.

### Face shield assembly

Following printing of the headband and bottom bracket, and laser cutting of the foam pad and the transparent visor, the face shield was assembled using the instructions in **Supplementary Material 3**. Briefly, the foam pad was attached to the inner band of the headband using either super glue or hot glue (unknown manufacturers), hook and loop straps (VELCRO Brand ONE-WRAP Double Sided Roll 0.75 in) were cut to 330 mm in length and secured to the headband by looping the hook and loop straps inside the hole at the posterior side of the headband, and then attaching the straps onto itself. The transparent visor was then mounted onto the headband by first securing one of the outer holes of the visor onto the headband peg. The visor was pulled across the headband so that each visor hole was aligned with the pegs of the headband. Prior to delivery for testing, face shields were cleaned using sanitizing wipes (Super Sani cloth, EPA registration number 9480-4) and placed under 254 nm ultraviolet light for 5 min in a germicidal cabinet (Monitor 2000, Sellstrom).

### Testing and Validation in Clinical Setting Subject Selection

To assess face shield usability and safety, a cohort of physicians, physician assistants, emergency department technicians, environmental service staff, and other individuals with patient-facing roles were recruited to the study from the BWH Emergency Department. To account for different workflows and preferences, participants were recruited from both day and night shifts. Study subjects were provided with a fact sheet and verbal consent was obtained (Partners Healthcare IRB: 2020P00910, **Supplementary Material 1**).

### Quality assessments

To assess quality, fabrication staff performed the following assessments in accordance with testing procedures reported for existing face shield designs^*8*^. (1) Visually inspected each component, checking for printing defects, cracks, and crevices. (2) Donned and doffed the face shield 10 times. Donning and doffing of the face shields were done in accordance with CDC guidelines.

### Functionality assessments

To assess functionality, research subjects were fitted with an unused PanFab face shield and the following tests were performed: (1) Test of splash resistance: a spray of water was delivered using a spray at the center of the visor. The visor passed the test if a subject did not feel any droplets on her/his face or neck. (2) Wearability testing: With the face shield on, subjects were asked to look left, right, up, down, and shake their heads, say yes and no. The face shield passed the test if none of the motions were impeded and the face shield did not fall off.

Fogging testing: The face shield was worn with and without a facemask for an extended period (min. 30 mins) under physical stress (e.g. an exercise machine) by one participant and it was not observed to undergo excessive fogging.

### User Feedback

An initial survey was administered to evaluate baseline demographics and attitudes towards PPE. After fit and splash testing, subjects returned to their work and used the face shield during their regular workflow for one hour, at which time a second survey was administered to obtain feedback on face shield performance.

## Results

### Description of design and iterative clinical feedback

We aimed to develop a locally fabricated face shield that would meet the requirements of an academic hospital when traditional supply chains failed. We produced a simple design that limited aerosol and splatter exposure coming from the front and above, that was resistant to fogging, and that was comfortable enough to be worn all day by healthcare professionals in a high-intensity clinical setting. Conventional disposable face shields on the US market are commonly available in 3/4 length (178 mm; 7 in.) and full length (230 mm; 9 in.) versions. We produced a full length visor, but increased its width from 230mm to 305mm so as to maximize face protection without obstructing hearing or impeding a user’s range of motion^3,9^. We produced a face shield that could be reused by a single individual following cleaning and disinfection procedures recommended by the CDC for reprocessing protective eyewear. Each face shield was used by only one user and was cleaned between uses with EPA-registered sanitizing wipes (Super Sani cloth, EPA registration number 9480-4; **Figure 1**).

**Figure 1:**
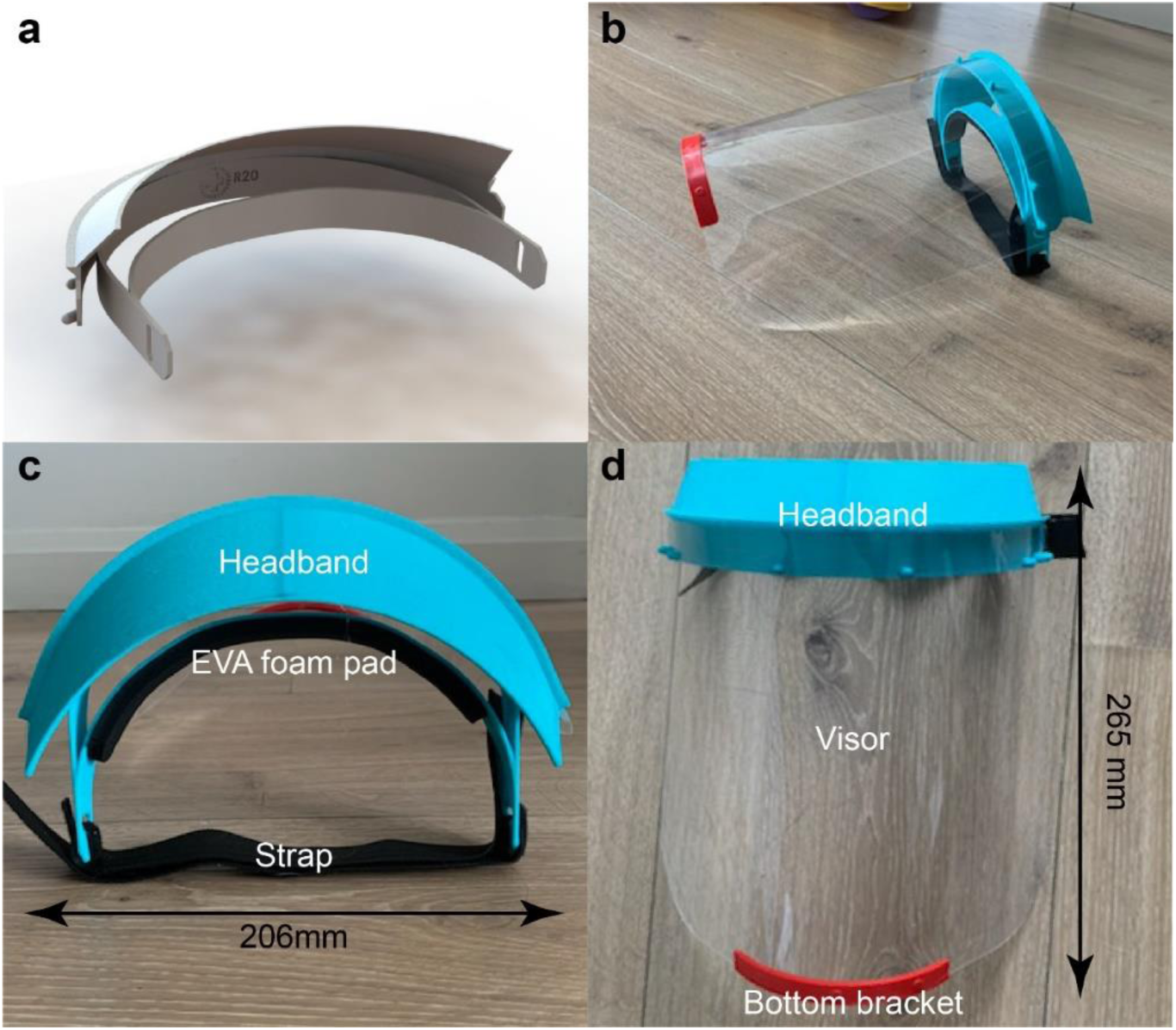
**A)** Headband CAD image **B)** final face shield prototype **C)** headband, foam pad, and strap image with dimensions **D)** headband, visor, and bottom bracket image with dimensions.

Starting with the Prusa RCX design (**Figure 2A**),^6^ we made iterative modifications based on clinician feedback and user testing. Our design, shown in **Figure 2B**, is similar in many respects to the DtM-v3.1 face shield which was subsequently released via the NIH 3D Print Exchange^8^. The similarity between the DtM-v3.1and the PanFab design is a result of convergence on a core set of features, the consistency of feedback across health care staff, and the applicability of the design to other health care settings. Expert user feedback was an essential part of the process. For example, potential users were concerned that the original Prusa design did not provide adequate liquid protection at the top and sides of the visor. We therefore added a fin above the headband to prevent fluid from entering the top of the face shield during high-risk procedures in which a clinician is required to lean forward; this includes endotracheal intubation for mechanical ventilation, one of the riskier procedures that must be performed on COVID-19 patients. We also added a lip above the visor so that any liquid that did fall on the fin would be retained by the lip and would not spread over the visor and affect a user’s ability to see through the face shield. Overall, four substantial design modifications were made based on clinical feedback, as outlined in **Table 2**. We note that the resulting design includes many features that are absent from disposable commercial face shields.

**Figure 2:**
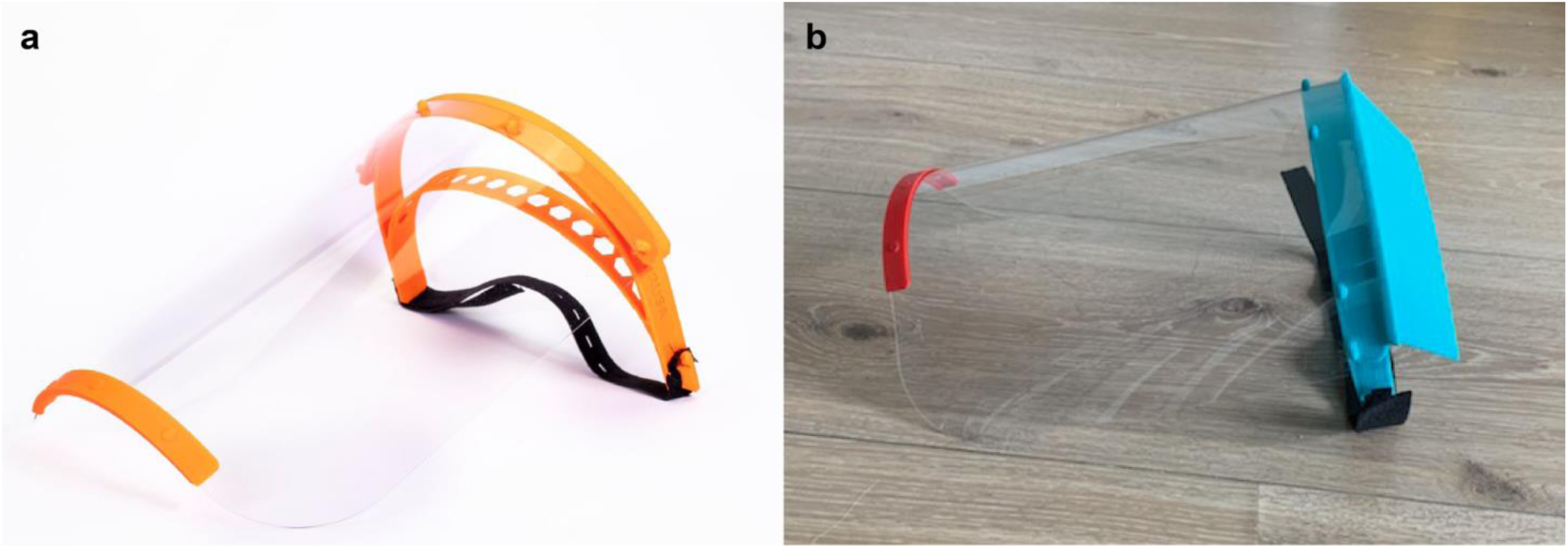
**A)** Image of Prusa design^6^ and **B)** final PanFab face shield prototype.

### Testing in a clinical environment

A total of 97 adults (≥18 years of age) in a variety of clinical roles at the BWH main campus Emergency Department were enrolled in the study. Five participants were lost to follow-up and were excluded from the analysis. Enrollment occurred during two shifts (daytime [n=52] and overnight [n = 40]) to account for potentially varied attitudes, patient volume, available resources, or other confounders. Demographic information and roles are summarized in **Table 3**. As described in the Methods, all study participants passed splash and fit tests before using the face shield in their typical duties.

**Table 3:** Demographics (Total Respondents: 92)

| Feature | Number | Percent |
| --- | --- | --- |
| **Sex** |  |  |
| Male | 25 | 27.2% |
| Female | 67 | 72.8% |
| **Size** |  |  |
| Mean Height (inches) |  | 66.2 |
| Mean Weight (lbs) |  | 164.3 |
| **Role** |  |  |
| Attending | 4 | 4.3% |
| Resident | 4 | 4.3% |
| Nurse | 45 | 48.9% |
| Tech | 16 | 17.4% |
| Physician Assistant | 6 | 6.5% |
| Environmental | 6 | 6.5% |
| Registration | 2 | 2.2% |
| Radiology | 5 | 5.4% |
| Other | 4 | 4.3% |

### Baseline Experiences with COVID and Attitudes on PPE

Each subject completed a questionnaire on baseline experiences and attitudes. The great majority of study subjects (81.4%) identified themselves as having a patient-facing, clinical role (e.g., physician, physician assistant, nurse, or technician); similarly, most (n = 88, 96%) reported having recently been directly involved in the care of a person under investigation (PUI) for possible coronavirus infection. Nearly all subjects had recently worn some form of eye protection (n = 91, 99%) and most had exclusively used personal protective equipment (PPE) that was hospital standard-issue (n = 57, 62%). Most respondents trusted hospital standard-issue PPE (n = 70, 76%), although some reported being unsure (n = 12, 13%) (**Table 4**).

**Table 4:** Baseline Experience and Attitudes

|  | Ever been involved in the care of a person with suspected COVID |  | Worn eye protection in the past week |  | Used non-hospital supplied PPE |  | Trust hospital supplied PPE |  |
| --- | --- | --- | --- | --- | --- | --- | --- | --- |
| Answer | Number | Percent | Number | Percent | Number | Percent | Number | Percent |
| Yes | 88 | 95.7% | 91 | 98.9% | 35 | 38.0% | 70 | 76.1% |
| No | 2 | 2.2% | 1 | 1.1% | 57 | 62.0% | 10 | 10.9% |
| Unsure | 2 | 2.2% | 0 | 0.0% | 0 | 0.0% | 12 | 13.0% |
| Total | 92 | 100.0% | 92 | 100.0% | 92 | 100.0% | 92 | 100.0% |

### Experience with reusable 3D-printed face shield in comparison to hospital standard-issue model

No respondents reported that the PanFab face shield was worse than the hospital standard-issue model in splash protection, durability, ease of use, or comfort; in fact, many preferred it over the hospital-issued model. Average scores in each of four categories (on a 5-point Likert scale) for splash protection, durability, ease of use, and comfort were 4.7, 4.6, 4.3 and 4.4, respectively (**Table 5**). This indicates a better experience with the PanFab face shield as compared to the hospital standard-issue model in all surveyed categories. Most participants rated the novel face shield as offering *slightly better* or *much better* splash protection (n = 87, 95%) and durability (n = 84, 91%, Table 5). Nearly all participants reported feeling *comfortable* or *very comfortable* using this face shield (n = 88, 96%), with only 1 person (1 %) stating she/he felt *neither comfortable nor uncomfortable* using the face shield (**Table 6)**. With respect to continued use, 92% (n = 85) of users planned to continue using the PanFab face shield; four respondents reported being unsure about continued use, but none were opposed (**Table 5**).

**Table 5.** Response across domains to the question: “compared to the standard issue face shield, how would you rate the prototype face shield?”

| Response* | Criterion (number of users) |  |  |  |
| --- | --- | --- | --- | --- |
|  | Comfort level with splash protection | Sturdiness and reliability | Ease of Use | Comfort |
| Much Worse | 0 | 0 | 0 | 0 |
| Slightly Worse | 0 | 2 | 3 | 5 |
| Not Worse/Not Better | 4 | 5 | 17 | 10 |
| Slightly Better | 16 | 17 | 21 | 17 |
| Much Better | 71 | 67 | 48 | 55 |
| **Average Score\*** | **4.7** | **4.6** | **4.3** | **4.4** |

\* Individual scores starting at 1 for “much worse” and extending to 5 for “much better”

**Table 6:** How comfortable are you using this shield in a clinical scenario where you did not have another option?

| Response | Number | Percent |
| --- | --- | --- |
| Very Uncomfortable | 0 | 0.0% |
| Uncomfortable | 0 | 0.0% |
| Neither Comfortable nor Uncomfortable | 1 | 1.1% |
| Comfortable | 27 | 30.3% |
| Very Comfortable | 61 | 68.5% |

### Participant comments

Anonymous respondent comments were also collated. Many expressed gratitude and thanks for the opportunity to use the PanFab face shield and felt that our efforts demonstrated support for frontline clinicians. The impact on morale is a positive aspect of community-resourced PPE, particularly when health care workers are under extremely difficult workplace conditions. Participants’ anonymous verbatim comments included “*I prefer these [new] shields to our old shields*”, “*This is very sturdy, comfortable and it doesn’t fog! [*…*] I plan to wear this every day*”, and “*[The] area of protection is amazing, feels sturdy and secure to head. [While there is] mild pressure on [the] forehead, [this is] preferable to [a different shield] that offers less protection*”. Other feedback included concerns that the Velcro strap might be a problem for some users with longer hair and that the shield length could be an issue for shorter users. These are issues we intend to address with additional design improvements.

## Discussion

This project highlights the ability of a voluntary collaboration involving designers, engineers, material scientists, clinicians, local fabricators and concerned citizens to perform rapid-cycle iterative prototyping developing 3D-printed PPE that addresses severe shortages in a time of crisis. The resulting BWH/PanFab Mk 1.0 face shield provides protection from spray in a reusable design that can be cleaned using standard hospital disinfectants. Using a team that self-organized on-line in response to a request of hospital incident command, PanFab was able to proceed from project inception to implementation in three weeks. Critically, using a clinical testing approach, we were able to introduce a non-traditionally manufactured product into a hospital supply chain. To date, we have fabricated hundreds of face shields, all of which remain in use, and we expect to introduce an additional thousand face shields within the coming weeks. Keys to successful integration of this face shield in the hospital setting included a dedicated liaison within incident command, the willingness of the IRB to work closely and quickly with designers, and the ability of the BWH legal and leadership teams to act quickly on policy issues. With a design in hand, we expect that others can deploy the solution we describe in as little as two weeks, plus 2-3 days for clinical testing (if required). In some cases, teams seeking to replicate our approach will need to modify the PanFab design due to shortages of raw materials or differences in fabrication capabilities. Under these circumstances, additional user testing under an IRB protocol might be required.

We are currently pursing three follow-on activities. First, we are developing designs that will make it possible to use injection molding as an alternative to 3D printing (see below). Second, we intend to subject our final design (printed or molded) to testing and certification under ANSI/ISEA Z.87.1-2015. We note however that design and testing standards are not freely available (they currently cost several hundred USD) and testing represents a substantial additional expense. Nonetheless, having a set of tested and approved designs would increase resiliency during a public health emergency such as COVID-19. Finally, we are testing various procedures for decontaminating and reusing face shields and similar PPE. Preliminary results with ionized hydrogen peroxide sterilization (iHP; TOMI SteraMist™)^10^ suggests that PanFab face shields can be successfully sterilized without suffering damage through at least five cycles.

### Additional considerations for large-scale manufacturing and dissemination

Moving from prototyping to large-scale manufacturing is a process that traditionally takes many months but in the context of a pandemic needs to be completed in a matter of weeks. 3D-printing and laser cutting are efficient methods for prototyping a design and improving it iteratively, but they are not ideal for large scale manufacturing. Alternative approaches include rotary die cutting to produce the face shield transparent visors and injection molding to fabricate the headband and support bracket. The transition from laser-cutting to rotary die cutting is straightforward, but injection molding the headband will require several adjustments to the design. This includes subtly changing the shapes of specific elements and selecting appropriate materials. Once complete, engineers can leverage the expertise of a company specialized in rapid-turnaround injection molding (e.g. Protolabs, Xcentric Mold in the US and similar companies in Europe and Asia) and quickly reach larger-scale production. In addition to enabling higher volume production than 3D printing, injection molding creates a more consistent product that is more likely to pass ANSI/ISEA testing. Injection molded parts can also be sterilized using a range of technologies whereas concerns have been raised about sterilization of 3D FFF parts made from PLA.^11^

### Regulatory considerations and proposed improvements

In collaboration with our local IRB, we continue to study how best to approach the problem of introducing an untested device, even one as simple as a face shield, into a hospital environment. In the current crisis, face shields are a likely prelude to testing and introduction of products in which safety considerations are more critical such as ventilator splitters. An IRB-approved protocol was used in the current study because we performed a user survey. However, use of a research protocol in this setting may have wider applicability if we consider a non-traditionally manufactured face shield as an Investigational Device. For *non-significant risk devices* such as face shields, the FDA authorizes IRBs to conduct the necessary risk assessment, and an Investigational Device Exemption (IDE) is not likely to be required from the FDA. We note however, that the relevant US regulations in 21 CFR 812.2 do not cover circumstances in which a normally approved device (i.e., a face shield meeting ANSI/ISEA Z.87.1-2015) that has become unavailable might have a non-approved variant (i.e., the PanFab face shield) that would be tested via research protocol. In the specific case of our deployment of the PanFab face shield, the latest emergency guidance from the FDA^5^ would appear to apply; some US state governments have issued their own guidance^12^.

Existing emergency guidance is not necessarily adequate for all anticipated needs in the current COVID-19 epidemic and it is neither guaranteed nor permanent. Some countries, such as Canada have more restrictive policies in place.^13^ Thus, we believe that it would highly desirable to establish procedures whereby IRB (or ethics committee) - approved research protocols could be used to facilitate future responses to medical emergencies and also promote much needed innovation in PPE. This regulatory clarification should specifically cover circumstances likely to arise in pandemic emergencies, when local fabrication is needed to augment strained supply chains.

When an emergency is over, devices that have not met prevailing regulatory requirements will likely need to be withdrawn from service to prevent continued use of products with unknown durability and performance characteristics. Precisely when and how this should occur remains unclear. Is it ethical for a hospital that no longer needs products made under emergency conditions to destroy them if other hospitals are in need? Conversely, is it ethical or legal to transfer unused unapproved products, or used but sterilized products? These issues remain largely unexplored.

### Lessons Learned

The global pandemic has put extreme pressure on health care systems and highlighted many weaknesses in the highly centralized supply chains that have developed for critical medical supplies. Local manufacturing represents an alternative source of supply in an emergency that has potential to rapidly address these shortages. Community-level disaster resilience is well-recognized as essential in responses to both natural disasters and public health emergencies,^14^ but the role of local manufacturing and maker communities in medical supply chains has not previously been considered part of such resiliency. We strongly believe that this should change and that such change will require refinement of regulatory and institutional policies. Hospitals should integrate individuals with engineering and manufacturing expertise into their incident command structure and prioritize longitudinal relationships with the local fabrication and maker communities well before an emergency happens. Our experience highlights the fact that individuals with the necessary medical, engineering and managerial experience already exist in many academic medical centers; such individuals need to be included in future pandemic planning.

The creation of research protocols for PPE testing could also bring much needed innovation in normal times. Studies over a period of at least 15 years by the US National Academies of Sciences and other US government bodies^15^ have repeatedly highlighted the need for innovation in PPE but little has changed. Practitioners and ordinary citizens should demand a much more transparent and distributed system for providing essential medical products of all types. Designs for key products should be tested clinically and published in peer-reviewed journals and demonstrated to meet existing fabrication standards well in advance. Unpatented designs for essential medical products should be made publicly available under non-restrictive Creative Commons or similar licenses. Patented designs should be placed in a patent pool for free use during public health emergencies or be subject to compulsory licensing at a reasonable cost. National suppliers and local fabricators must be compensated for their work but in extreme cases, 28 U.S. Code § 1498 (“Section 1498”) gives the US Federal government the “right to use patented inventions without permission, while paying the patent holder ‘reasonable and entire compensation’,” with immunity from patent claims. The current crisis has shown that, when a pandemic is spreading, and health care workers are placed at high risk, we require a distributed and robust *community level* approach to essential medical supplies, not a secretive and centralized one. The resulting devices, developed and produced largely by volunteers, are not only likely to decrease the risk of hospital infection in the current example, but also send a powerful message to front-line medical staff that the local community stands behind them. Although a pandemic was required to galvanize these insights and promote rapid change, our hope is that the spirit of thoughtful collaboration and rapid innovation does not dissipate once the world returns to its pre-COVID-19 state.

## Data Availability

The designs generated during this study are included in this published article (and its supplementary information files). The datasets generated during and/or analysed during the current study are available from the corresponding author on reasonable request.

## Acknowledgements

Above all we thank the members of the Greater Boston Pandemic Fabrication Team (PanFab) for technical, administrative, and logistic support necessary for the execution of this project. Membership found at https://www.panfab.org/the-team-and-the-project/consortium-members. We also thank Michael Klompas, MD, Jon Boyer, ScD CIH, and Charles M. Morris, MD for their clinical and safety feedback, Kevin T. Giordano, MBA FACHE, Douglas Carney, AIA MBA, Julia Sinclair, MBA, Allison Moriarty, MPH, Bernard R. Jones, EdM for their support with implementation, operations and approval process, Peter Chai MD for helping with protocol development, as well as all of the healthcare workers that participated in the study.

## Author Contributions

Face shield design prototyping and production: M.J.A., B.B, P.D.A., E.B., A.F., J.F., R.O., L.S, C.V., S.H.Y. Face shield design clinical testing: A.M., P.D.A., N.L., S.H.Y. Writing: A.M.,.M.J.A, D.P., M.S.S., P.K.S., N.L., S.H.Y. Greater Boston Pandemic Fabrication Team (PanFab) Consortium Coordination: D.P., H.Y., P.K.S.

## Outside interests

A Mostaghimi is a consultant or has received honoraria from Pfizer, 3Derm, and hims and has equity in Lucid Dermatology and hims. He is an associate editor for JAMA Dermatology. Mostaghimi declares that none of these relationships are directly or indirectly related to the content of this manuscript.

PK Sorger is a member of the SAB or Board of Directors of Applied Biomath, Glencoe Software and RareCyte Inc and has equity in these companies. In the last five years the Sorger lab has received research funding from Novartis and Merck. Sorger declares that none of these relationships are directly or indirectly related to the content of this manuscript.

NR LeBoeuf is a consultant for or has received honoraria from the following companies:Seattle Genetics, Sanofi and Bayer

Edward W Boyer is funded by NIH grants R01DA047236, HL R01HL 126911, HD093655, DARPA award FA8750-18-C-0025, and Philips Healthcare. Boyer declares that none of these relationships are directly or indirectly related to the content of this manuscript

Philip D Anderson has equity in Hallandia V. Anderson declares that none of these relationships are directly or indirectly related to the content of this manuscript.

## Funding Information

Local fabricators, makers and citizens generously donated their time and resources and were essential for all stages of the project. This work was also supported by the Harvard MIT Center for Regulatory Sciences and by NIH/NCI grants U54-CA225088 (to PKS, NL and DP) and by T32-GM007753 (to DP) and by the Harvard Ludwig Center. MJA is a recipient of the Friends of McGovern Graduate Fellowship.

## Supplementary Materials

Supplementary material 1: Associated study IRB questionnaire

Supplementary material 2: Design files of face shield parts: .STL + DXF

Supplementary material 3: Instruction for use and overview of product

## Notes

### Competing Interest Statement

The authors have declared no competing interest.

